# Utility of Automated Data-adaptive Propensity Score Method for Confounding by Indication in Comparative Effectiveness Study in Real World Medicare and Registry Data

**DOI:** 10.1101/2021.04.06.21254887

**Authors:** Hiraku Kumamaru, Jessica J. Jalbert, Louis L. Nguyen, Lauren A Williams, Hiroaki Miyata, Soko Setoguchi

## Abstract

**Background:** Confounding by indication is a serious threat to comparative studies using real world data. We assessed the utility of automated data-adaptive analytic approach for confounding adjustment when both claims and clinical registry data are available.

**Methods:** We used a comparative study example of carotid artery stenting (CAS) vs. carotid endarterectomy (CEA) in 2005-2008 when CAS was only indicated for patients with high surgical risk. We included Medicare beneficiaries linked to the Society for Vascular Surgery’s Vascular Registry >65 years old undergoing CAS/CEA. We compared hazard ratios (HRs) for death while adjusting for confounding by combining various 1) Propensity score (PS) modeling strategies (investigator-specified [IS-PS] vs. automated data-adaptive [ada-PS]); 2) data sources (claims-only, registry-only and claims-plus-registry); and 3) PS adjustment approaches (matching vs. quintiles-adjustment with/without trimming). An HR of 1.0 was a benchmark effect estimate based on CREST trial.

**Results:** The cohort included 1,999 CAS and 3,255 CEA patients (mean age 76). CAS patients were more likely symptomatic and at high surgical risk, and experienced higher mortality (crude HR=1.82 for CAS vs. CEA). HRs from PS-quintile adjustment without trimming were 1.48 and 1.52 for claims-only IS-PS and ada-PS, 1.51 and 1.42 for registry-only IS-PS and ada-PS, and 1.34 and 1.23 for claims-plus-registry IS-PS and ada-PS, respectively. Estimates from other PS adjustment approaches showed similar patterns.

**Conclusions:** In a comparative effectiveness study of CAS vs. CEA with severe confounding by indication, ada-PS performed better than IS-PS in general, but both claims and registry data were needed to adequately control for bias.

## INTRODUCTION

Real-world data (RWD) including large electronic databases such as claims databases, electronic health records databases, and patient/procedure registries are rich sources of data for conducting comparative effectiveness and safety studies. While these studies may be used to complement findings from randomized clinical trials for cardiovascular disease,^1–6^ confounding is one of the biggest challenges to study validity. Confounding by indication is particularly problematic when comparing the effectiveness of two or more alternative treatment modalities because each modality may be indicated for clinically distinct patient groups. For example, carotid artery stenting (CAS) was originally indicated only for patients with high surgical risk when its use was first approved for reimbursement by CMS in 2004^7^. It was not until 2011 when a guideline, and observational comparative effectiveness rerecommended CAS for non-high risk patients based on later trials such as CREST proving the efficacy in the non-high risk population^8^. Thus, comparative effectiveness studies of CAS vs. carotid endarterectomy (CEA) using early phase data would likely be confounded by indication.

Most claims databases lack detailed clinical information to fully account for confounding by indication, and adding clinical information through data linkage with registries has been shown to greatly improve the validity of comparative studies in patients with cardiovascular diseases^2,3^. At the same time, registries may not be available or linkage may not be possible. Various methodological developments such as propensity score (PS)-based data-adaptive methods with automated variable generation and selection algorithms have been proposed to improve the validity of results using RWD (^9–13^). However, in the presence of linked clinical information from a registry, the relative utility of such approaches vs. additional clinical information in CER studies with strong confounding by indication has not been fully elucidated.

The aim of the present study was to evaluate the performance of the automated data-adaptive PS approaches for confounding adjustment in a CER study with strong confounding by indication when claims data only, registry data only, and linked claims plus registry data are available. We used the example of CAS vs. CEA using data from 2005-2008 when CMS coverage indication for CAS were limited to high surgical risk patients with predefined levels of stenosis severity and symptoms^14^.

## METHODS

### Data Sources

We used the SVS-VR, 2005-2008 as well as the denominator, institutional, non-institutional, and vital status Medicare files (2000-2009). The design and protocols of SVS-VR have been described in detail elsewhere^15^. The SVS-VR collects detailed information on medical history, pre-procedural diagnostics (including degree of carotid stenosis, symptomatic status, and high surgical risk status), procedure-related factors, and intra-operative and pre-discharge complications for patients undergoing CAS as well as CEA, allowing for comparison of the two procedures. The SVS-VR data are audited to ensure that all cases of CAS and CEA are reported and to verify the accuracy and completeness of the data^15^.

From CMS, we obtained administrative claims data from January 1^st^, 2000 to December 31^st^, 2009 for patients who underwent CAS (ICD-9-CM codes: 00.61, 00.63, 00.64) or CEA (ICD-9-CM codes: 38.12) in an inpatient setting, including information on patient demographics, eligibility, inpatient and outpatient facility services use, physician services use covered under Part B, and date of death.

Linkage of SVS-VR to the Medicare data has been previously described^16^. Briefly, we deterministically linked CAS and CEA procedures in the registry to the Medicare institutional file using date of birth, sex, facility, and the requirement that the procedure date in the registry be between a hospital admission and discharge date for the same procedure in the Medicare data.

### Patients

The patient population included in this study has been described^17^. In short, the study sample included Medicare fee-for-service beneficiaries who had undergone CAS or CEA at SVS-VR-participating facilities between 2005 and 2008, who were at least 66 years of age at the time of the procedure, were eligible for Medicare for at least one year prior to the procedure, and for whom Medicare was the primary payer. If patients underwent multiple procedures, the first procedure was selected. We set the day of the procedure as the index date and the preceding 1-year period as the baseline period. We followed patients from the index date until the earliest occurrence of one of the following events: death from any cause, loss of Medicare eligibility, or the end of the study period (December 31^st^, 2009).

### Outcome

The outcome of interest was death from any cause following carotid revascularization over the study period.

### Study Variables

#### Medicare claims data

We extracted data on patient age, sex, and race from Medicare denominator files. We also derived measures of healthcare utilization, such as number of past-year hospitalizations, physician visits, and nursing home admissions^18^. Patients were categorized as having undergone elective CAS unless the hospital admission type was defined as urgent, emergent, or of a traumatic nature in the institutional Medicare file. Patients were categorized as having symptomatic carotid stenosis if they had recorded diagnoses of stroke, transient ischemic attack, or amaurosis fugax in the year leading up the procedure. We also defined comorbidities using the diagnoses and procedure codes recorded during the baseline period.

#### SVS-VR

Information on the ipsilateral and contralateral degrees of carotid stenosis was available from interpreted pre-procedural carotid ultrasound and angiogram exam results. We categorized patients as having mild (<50%), moderate (50-69%), or severe (≥70%) ipsilateral carotid stenosis or contralateral stenosis ≥70% using information from ultrasound results and, when ultrasound data were missing, from angiograms. We considered patients to be at high surgical risk if they met any of the following criteria: age >80, New York Heart Association (NYHA) class III or IV heart failure, left ventricular ejection fraction <30%, unstable angina, myocardial infarction in the past 30 days, recurrent stenosis, prior radical dissection or radiation, contralateral occlusion, contralateral laryngeal nerve palsy or injury, high anatomic lesion, or other physiologic or anatomic surgical risk factors from investigational device exemption trials of CAS. We classified patients as per the National Coverage Determination’s indications for CAS^14^: high surgical risk with symptomatic stenosis 50-69%, high surgical risk with symptomatic stenosis ≥70%, high surgical risk with asymptomatic carotid stenosis ≥80%, or those not matching the conditions above. We also obtained information on comorbidities from the registry.

### Propensity Score Modeling

We employed: 1) investigator-specified approach and 2) automated data-adaptive approach. For investigator-specified PS model, the investigators preselected the list of variables based on clinical knowledge and prior literature. For automated data-adaptive model, we chose high dimensional (hd) PS, one of the most commonly used algorithms.. Full lists of variables selected for each model are described in the footnote of Table 2.

**Table 1:**
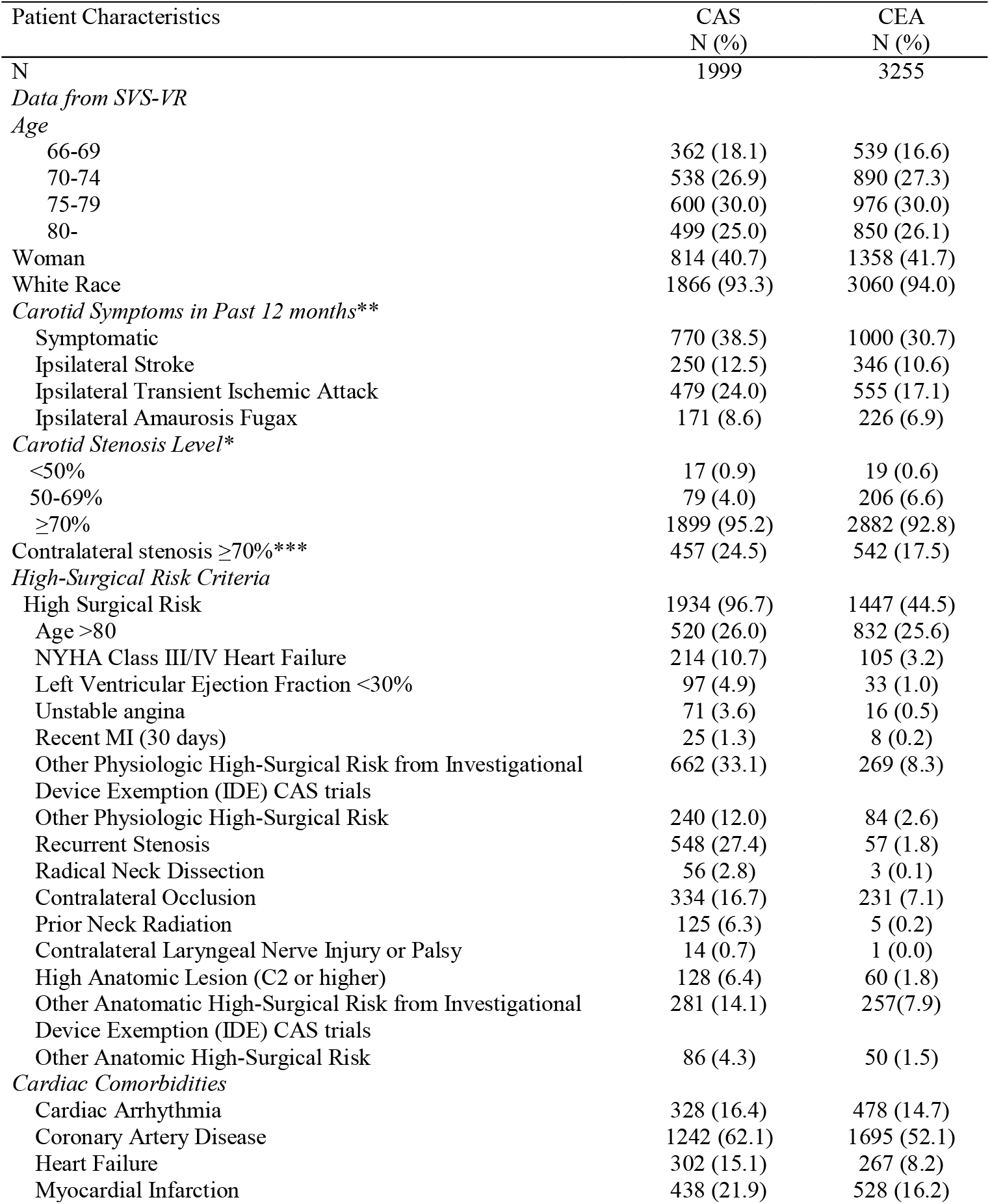
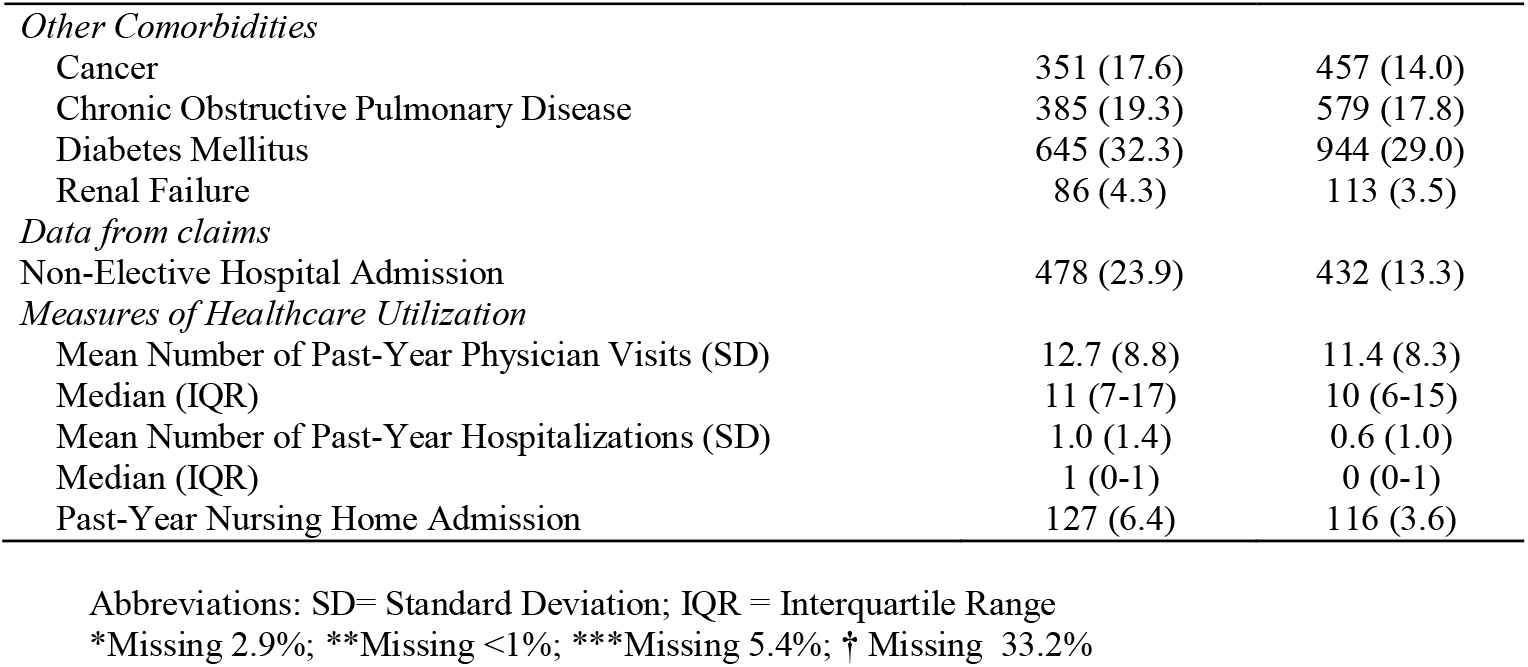
Selected Patient Characteristics at Baseline, By Procedure Patient Characteristics

**Table 2:**
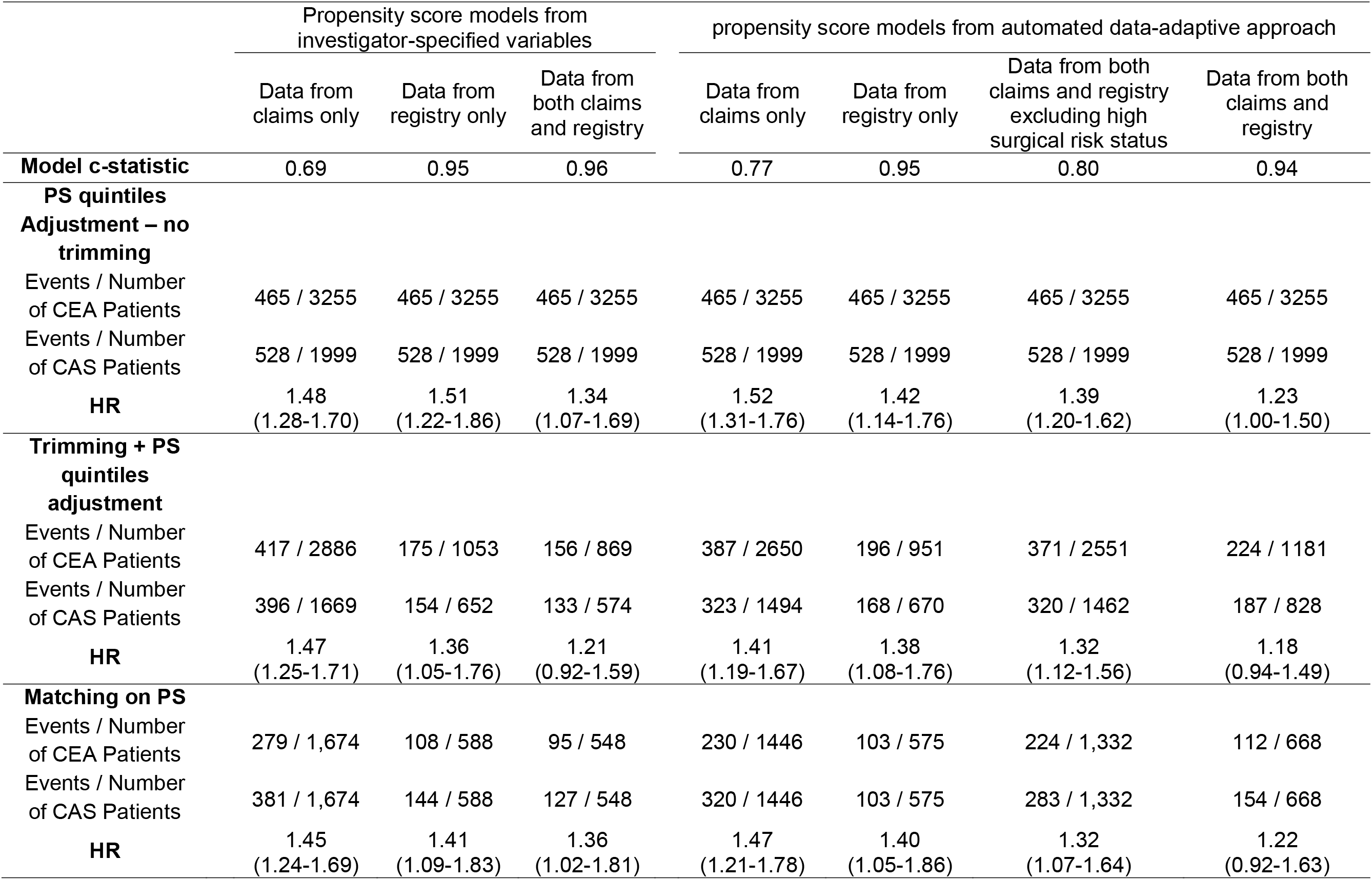
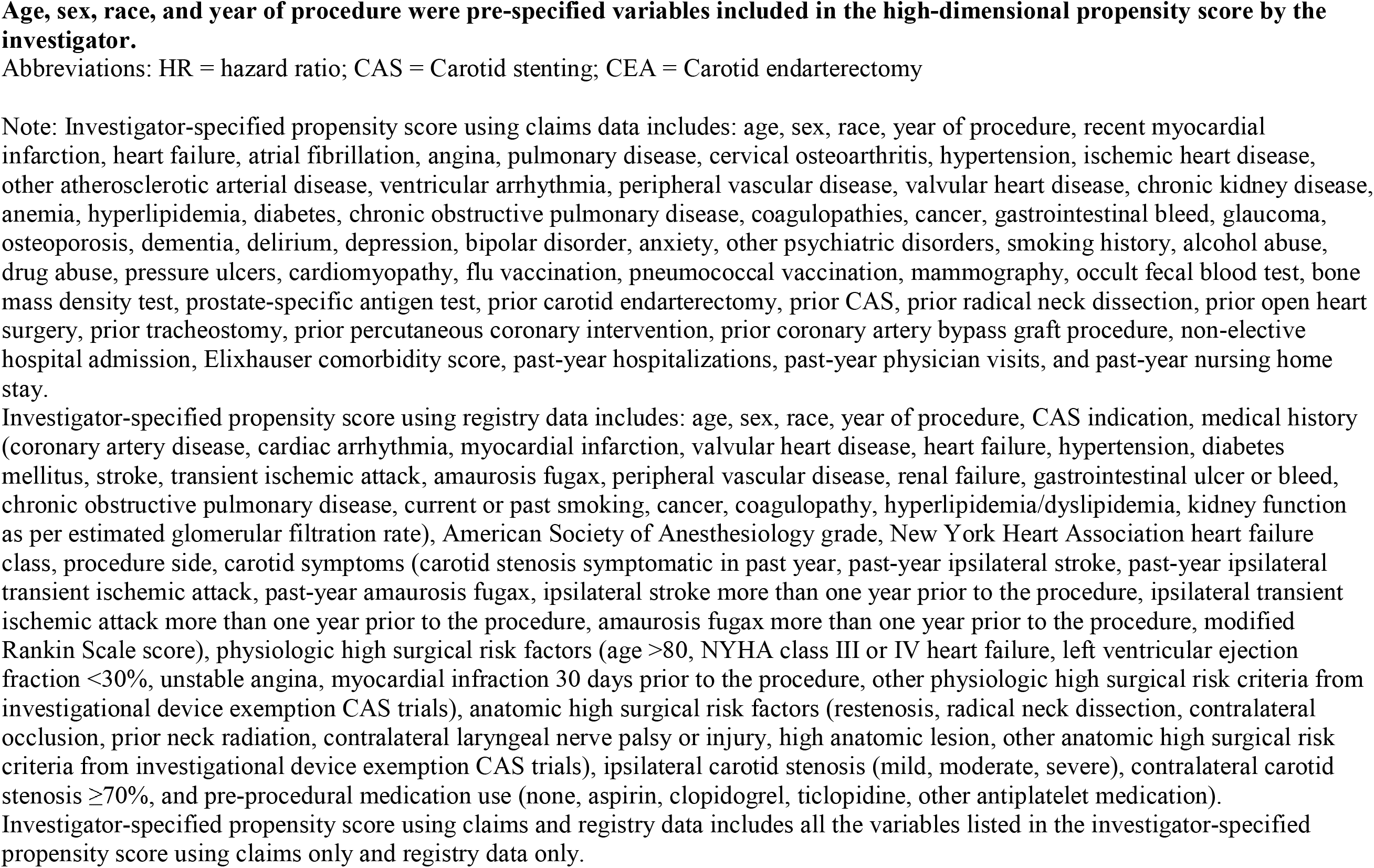
Number of Events, Total Cell Sizes and Hazard Ratios for CAS Relative to CEA Based on Different Propensity Score Adjustment Techniques Across the Different Propensity Score Models

**Table 3:**
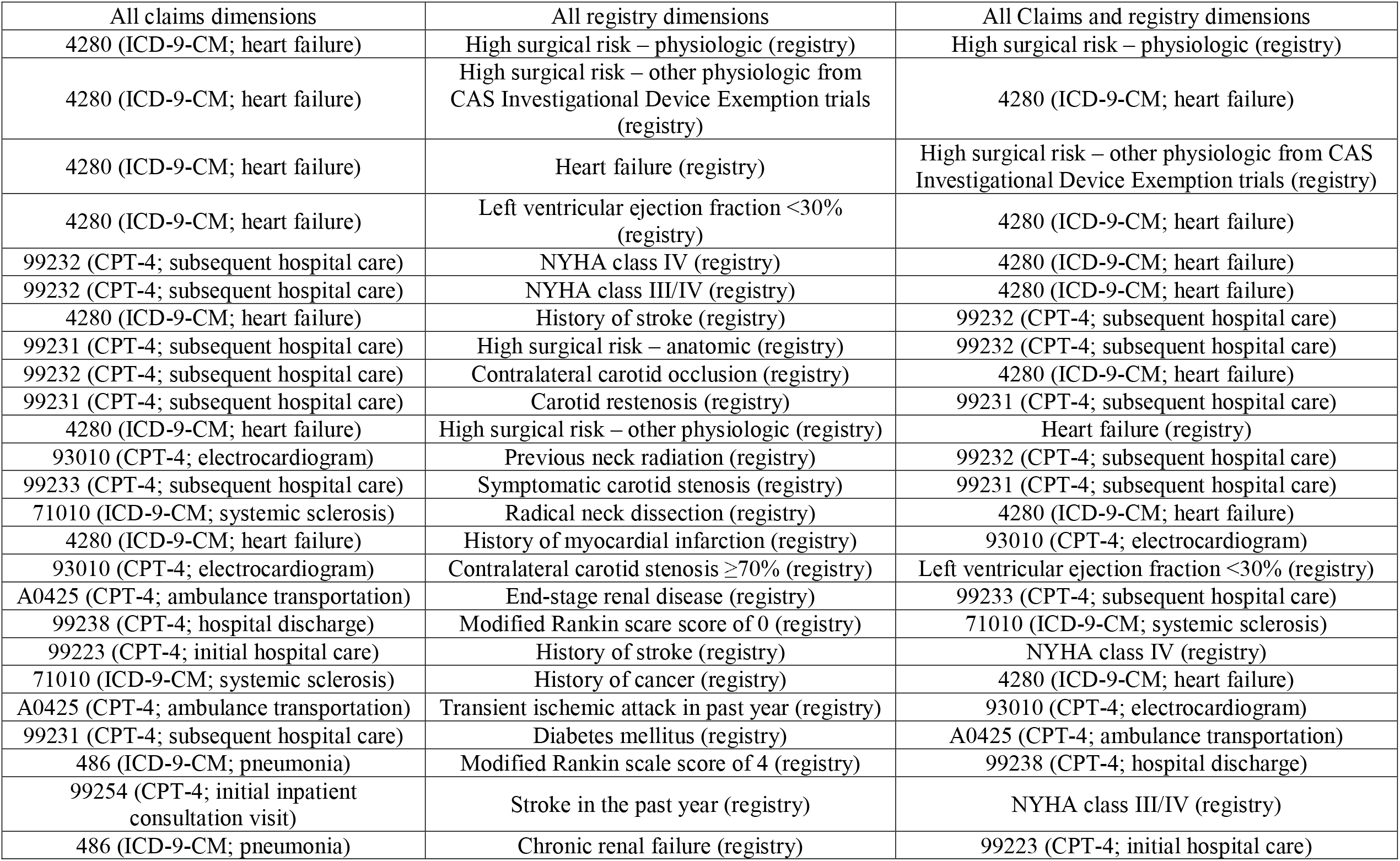
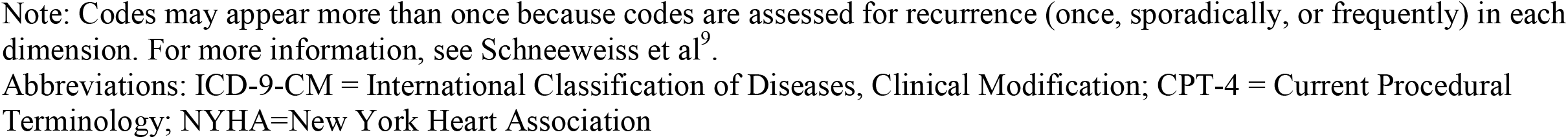
Top 25 Variables Identified by the High-Dimensional Propensity Score Algorithm as Having the Greatest Potential to Cause Confounding

| All claims dimensions | All registry dimensions | All Claims and registry dimensions |
| --- | --- | --- |
| 4280 (ICD-9-CM; heart failure) | High surgical risk – physiologic (registry) | High surgical risk – physiologic (registry) |
| 4280 (ICD-9-CM; heart failure) | High surgical risk – other physiologic from CAS Investigational Device Exemption trials (registry) | 4280 (ICD-9-CM; heart failure) |
| 4280 (ICD-9-CM; heart failure) | Heart failure (registry) | High surgical risk – other physiologic from CAS Investigational Device Exemption trials (registry) |
| 4280 (ICD-9-CM; heart failure) | Left ventricular ejection fraction <30% (registry) | 4280 (ICD-9-CM; heart failure) |
| 99232 (CPT-4; subsequent hospital care) | NYHA class IV (registry) | 4280 (ICD-9-CM; heart failure) |
| 99232 (CPT-4; subsequent hospital care) | NYHA class III/IV (registry) | 4280 (ICD-9-CM; heart failure) |
| 4280 (ICD-9-CM; heart failure) | History of stroke (registry) | 99232 (CPT-4; subsequent hospital care) |
| 99231 (CPT-4; subsequent hospital care) | High surgical risk – anatomic (registry) | 99232 (CPT-4; subsequent hospital care) |
| 99232 (CPT-4; subsequent hospital care) | Contralateral carotid occlusion (registry) | 4280 (ICD-9-CM; heart failure) |
| 99231 (CPT-4; subsequent hospital care) | Carotid restenosis (registry) | 99231 (CPT-4; subsequent hospital care) |
| 4280 (ICD-9-CM; heart failure) | High surgical risk – other physiologic (registry) | Heart failure (registry) |
| 93010 (CPT-4; electrocardiogram) | Previous neck radiation (registry) | 99232 (CPT-4; subsequent hospital care) |
| 99233 (CPT-4; subsequent hospital care) | Symptomatic carotid stenosis (registry) | 99231 (CPT-4; subsequent hospital care) |
| 71010 (ICD-9-CM; systemic sclerosis) | Radical neck dissection (registry) | 4280 (ICD-9-CM; heart failure) |
| 4280 (ICD-9-CM; heart failure) | History of myocardial infarction (registry) | 93010 (CPT-4; electrocardiogram) |
| 93010 (CPT-4; electrocardiogram) | Contralateral carotid stenosis $\geq 70\%$ (registry) | Left ventricular ejection fraction <30% (registry) |
| A0425 (CPT-4; ambulance transportation) | End-stage renal disease (registry) | 99233 (CPT-4; subsequent hospital care) |
| 99238 (CPT-4; hospital discharge) | Modified Rankin score of 0 (registry) | 71010 (ICD-9-CM; systemic sclerosis) |
| 99223 (CPT-4; initial hospital care) | History of stroke (registry) | NYHA class IV (registry) |
| 71010 (ICD-9-CM; systemic sclerosis) | History of cancer (registry) | 4280 (ICD-9-CM; heart failure) |
| A0425 (CPT-4; ambulance transportation) | Transient ischemic attack in past year (registry) | 93010 (CPT-4; electrocardiogram) |
| 99231 (CPT-4; subsequent hospital care) | Diabetes mellitus (registry) | A0425 (CPT-4; ambulance transportation) |
| 486 (ICD-9-CM; pneumonia) | Modified Rankin scale score of 4 (registry) | 99238 (CPT-4; hospital discharge) |
| 99254 (CPT-4; initial inpatient consultation visit) | Stroke in the past year (registry) | NYHA class III/IV (registry) |
| 486 (ICD-9-CM; pneumonia) | Chronic renal failure (registry) | 99223 (CPT-4; initial hospital care) |

Note: Codes may appear more than once because codes are assessed for recurrence (once, sporadically, or frequently) in each dimension. For more information, see Schneeweiss et al<sup>9</sup>.
Abbreviations: ICD-9-CM = International Classification of Diseases, Clinical Modification; CPT-4 = Current Procedural Terminology; NYHA=New York Heart Association

The hdPS algorithm empirically generates and selects covariates that collectively act as proxies for the patient’s health status from the database for inclusion in the propensity score modeling. The full description of this algorithm is available elsewhere^9,19^. Briefly, the algorithm consists of three parts: (1) empirical variable identification; (2) variable ranking (prioritization); and (3) variable selection. First, the algorithm goes through each type of data such as inpatient diagnosis, inpatient procedure, namely ‘data dimension’ and generates covariates that represent the presence or absence of each code meeting a prespecified prevalence threshold (e.g. 1% or more). Second, the generated variables are ranked based on its potential to cause confounding, measured by its association with the outcome and the exposure^17^. Third, top 500 (or the number the investigators select) of these empirically selected variables are included to develop a PS model. In our study, ‘data dimensions’ considered for hdPS included inpatient, outpatient, non-institutional (carrier), and nursing home diagnosis and procedure codes. We also categorized variables in the registry into registry-based ‘data dimensions’ including medical history, symptomatic status, diagnostic imaging results, pre-procedural medications, and high-surgical risk status.

We constructed three propensity score models in each of the investigator-specified and hdPS approach using data dimensions in claims data only, with all SVS-VR variables only, and with claims data and SVS-VR variables (Table 2). We also included an additional PS model (4^th^ model in hdPS approach) using both registry and claims data but excluding the information on patients’ high surgical risk status to assess the importance of the high surgical risk status in confounding adjustment (Table 2).

### Statistical Analysis

We evaluated the baseline characteristics of the study cohort undergoing CAS or CEA using both registry and claims data. We report crude cumulative risks and 95% confidence intervals for 3-year mortality, derived from Kaplan-Meier estimators. We used multiple imputation to maximize power and handle missing covariate information from the SVS-VR (less than 5% of data were missing for all variables except contralateral carotid occlusion [5.4%], modified Rankin Scale score [5.5%], creatinine level [9.8%], NYHA class [17.6%], and hyperlipidemia/dyslipidemia [33.2%]).

Hazard for all cause death following CAS compared to CEA were derived using Cox proportional hazards regression models. The propensity score was included in the Cox model in three ways: using 1:1 nearest-neighbor matching with a caliper of 0.2 times the logit of the propensity score, and grouping patients into propensity score quintiles with and without implementing 5% asymmetric trimming^20^. We used the sandwich variance estimator to account for clustering of patients at the physician and hospital levels. The hazard ratios from these models were compared to the benchmark risk ratio of 1.0 based on the findings reported in randomized clinical trials^21,22^. All analyses were performed using SAS version 9.2 (SAS Institute, Cary, NC). The study was approved by the Institutional Review Board of Brigham and Women’s Hospital.

## RESULTS

We successfully linked 2867 SVS-VR CAS records and 4381 SVS-VR CEA records to Medicare claims data^3^. After applying inclusion and exclusion criteria, 1999 CAS patients and 3255 CEA patients were included in the study cohort (Figure 1).

**Figure 1:**
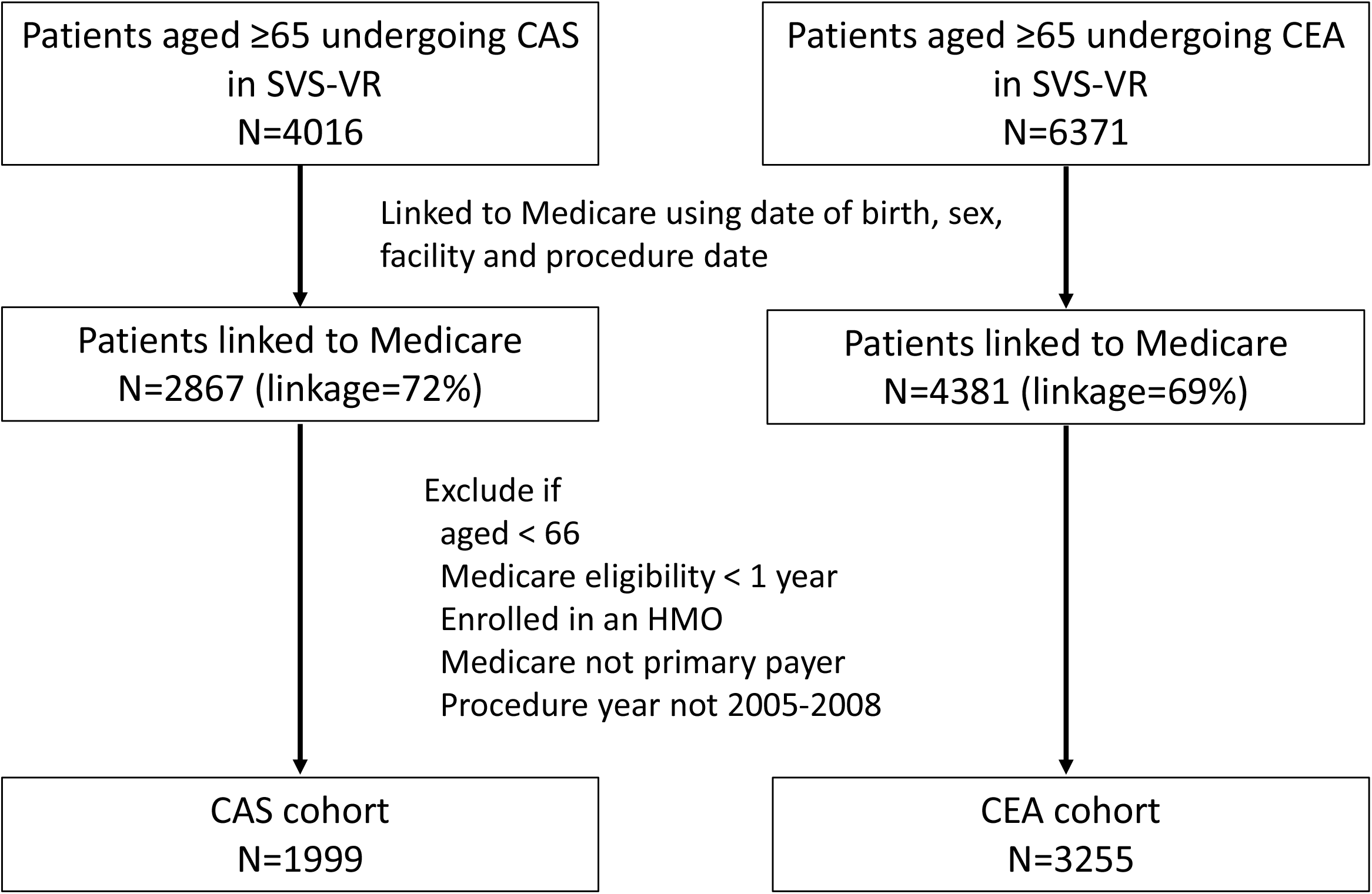
Flow Chart for Creation of Medicare-Linked SVS-VR CAS and CEA Study Cohorts. Abbreviations: CAS = Carotid stenting; SVS-VR = Society of Vascular Surgeons Vascular Registry; CEA = Carotid endarterectomy; HMO = Health maintenance organization;

The full characteristics of the population is described in the previous article by Jalbert and colleagues^17^. Briefly, the distribution of age, sex and race of the patients were similar between the two groups (Table 1). CAS patients were more likely to be symptomatic (CAS 38.5% vs. CEA 30.7%) and had a higher prevalence of high surgical risk factors (96.7% vs. 44.5%). Specifically, patients undergoing CAS had higher percentages of patients with class III/IV heart failure (10.7% vs. 3.2%), left ventricular ejection fraction < 30% (4.9% vs. 1.0%), unstable angina (3.6% vs. 0.5%), recurrent stenosis (27.4% vs. 1.8%), contralateral occlusion (16.7% vs. 7.1%), prior neck radiation (6.3% vs. 0.2%), and high anatomic lesion (6.4% vs. 1.8%) than patients undergoing CEA. CAS patients had a higher prevalence of cardiac comorbidities (e.g., coronary artery disease, 62.1% vs. 52.1%; myocardial infarction, 21.9% vs. 16.2%), and slightly higher prevalence of non-cardiac comorbidities including cancer (17.6% vs. 14.0%), COPD (19.3% vs. 17.8%), and renal failure (4.3% vs. 3.5%).

CAS patients were followed for a median of 909 days, during which time 446 deaths were identified (incidence of 3-year mortality: 25.5% (95% CI: 23.3-27.6)) (Figure 2). Similarly, we identified 424 deaths during 847 days of median follow-up (incidence of 3-year mortality: 16.8% (95% CI: 15.2-18.3)). The estimated crude hazard ratio was 1.82 (95% CI: 1.60-2.08).

**Figure 2.**
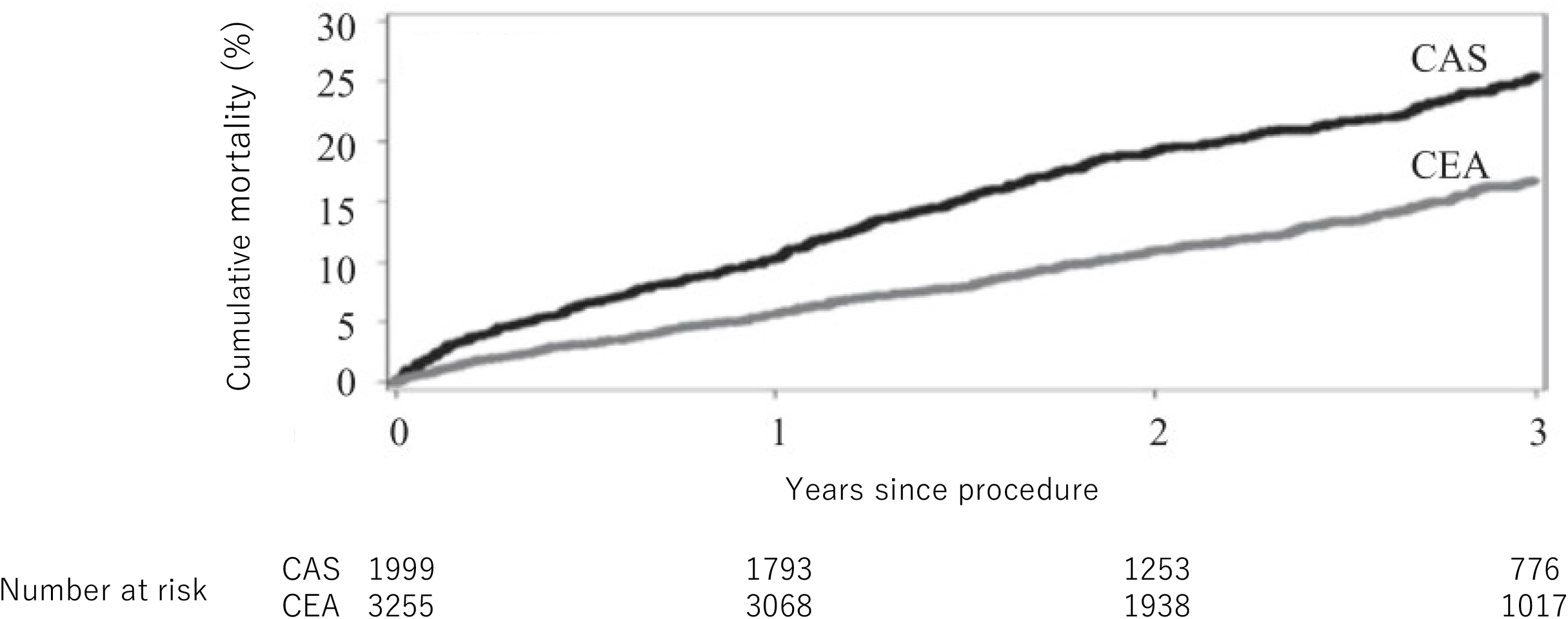
3-year cumulative incidence of death among the Medicare-linked SVR-VR patients, by procedure. Abbreviations: CAS=Carotid Stenting; CEA = Carotid endarterectomy

Figure 3 shows the distribution of the propensity scores from the six models with different combinations of data source and variable selection approach. All PS models that included registry data (A, C, D, F) clearly separated the distribution between the CAS group and the CEA group. The PS models using claims data only (B, E) resulted in much greater overlap between the two groups, indicating less ability to discriminate between those receiving CAS and those receiving CEA. The c-statistics of the propensity score models ranged from 0.69 to 0.96. The difference in the PS distributions and c-statistics for IS-PS and hdPS were identical or nearly identical except for the models using claims data only (B, E) where the discrimination between the two groups was higher using hdps (c-statistic of 0.77 vs. 0.69). Among the 4 models using either registry data alone or registry plus claims data (A,C,D,F), the c-statistics were similar regardless of whether investigator-specified variables or hdPS-selected variables were used. All models with information on surgical risk had c-statistics of 0.94 and above, while none without had such high c-statistics (≤0.80) (Table 2).

**Figure 3.**
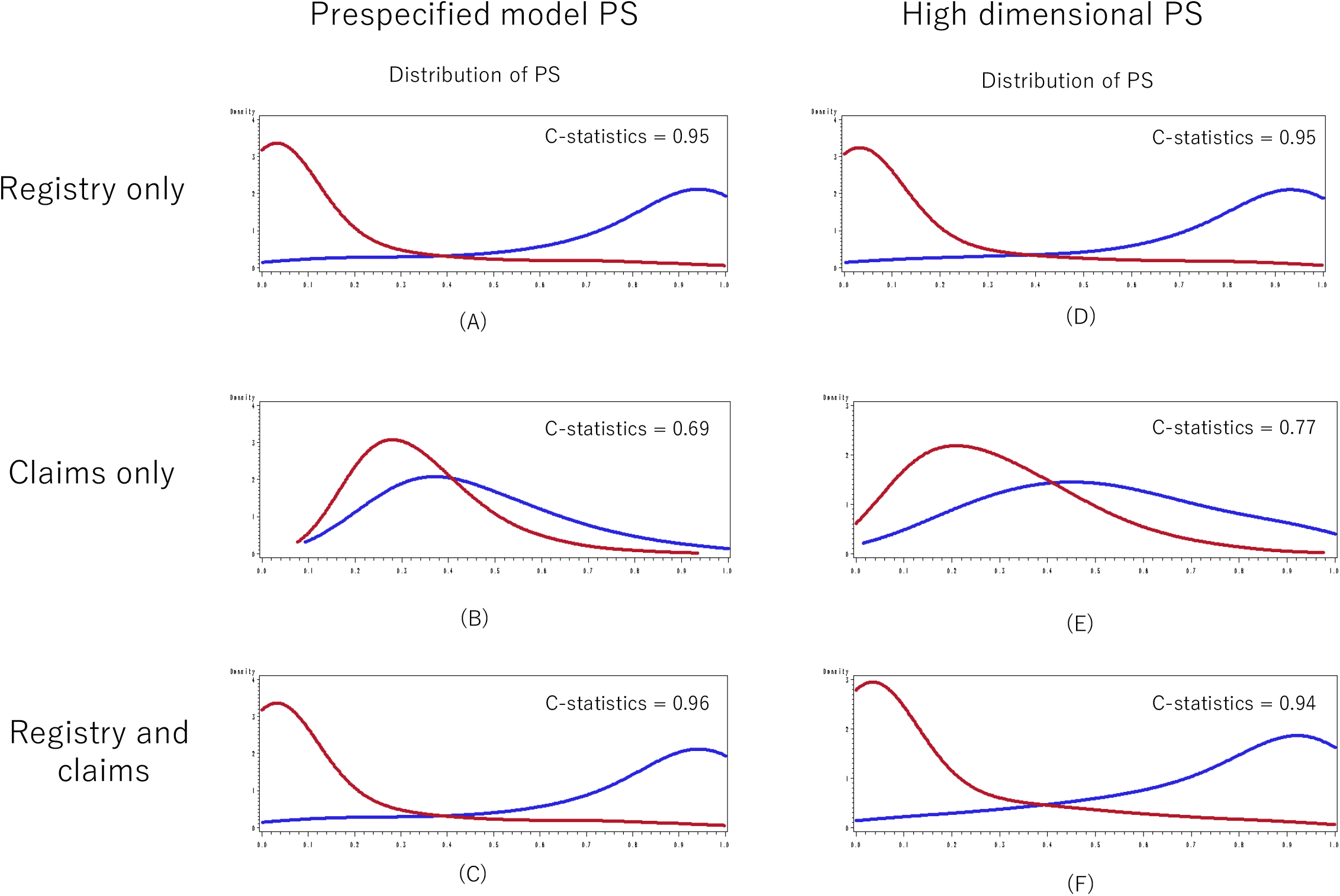
Distribution of the propensity scores estimated from the models with different combinations of data source and variable selection approach Abbreviations: PS = propensity score;

In the PS-matched analysis, the largest number of patients were retained when the investigator-specified PS model with claims data only was used (1674 patients each in the CAS and CEA groups). The adjusted HR estimate in the matched cohort was 1.45 (95% CI: 1.24-1.69), significantly different from the benchmark value of 1.0. The HR estimates were similar for the models using either registry data only or claims only, with or without the use of hdPS approach for variable selection. Investigator-specified propensity scores yielded a point estimate slightly closer to the benchmark of 1.0 at 1.36 (1.02-1.81) when both claims and registry data were used, and hdPS from claims plus registry data yielded a smaller HR estimate of 1.22 (0.92-1.63), which was the only estimate without statistical significance across the different approaches compared. The hdPS model with exclusion of high surgical risk status yielded an HR estimate of 1.32 (1.07-1.64) that was further away from the benchmark.

The findings from the two PS adjustment approaches were similar to those from the matched analysis. PS quintile adjustment without trimming yielded HR estimates slightly larger (i.e., away from the benchmark) compared to the matched analyses, although the difference was small. On the other hand, PS quintile adjustment with asymmetric trimming yielded point estimates that were generally similar or slightly smaller than those from the corresponding matched analyses, despite having more patients in the cohort than the matched analyses. In both adjustment approaches, hdPS from claims plus registry data yielded the smallest estimated HR; hdPS without information on high surgical risk status yielded an HR further from the benchmark.

The top 25 variables identified by the hdPS algorithm as having the greatest potential to cause confounding from claims data, registry data and the combination of the two are listed in Table 2. The variables selected from registry data were clinical factors, most of which matched the investigator-specified PS variables. Variables selected from claims data included clinical factors such as diagnoses for heart failure and pneumonia, as well as CPT codes indicating health services use such as hospital care, electrocardiogram, and ambulance transportation. The factors selected via the hdPS algorithm when both registry data and claims data were combined were a mix of these two.

## DISCUSSION

In a cardiovascular CER study with severe confounding by indication, we compared the performance of various methods for confounding adjustment using propensity scores (both investigator-specified and automated data-adaptive method) and using both claims and registry data. The c-statistics for the propensity score models were high and were similar for models from registry data with or without claims. However, propensity scores from claims data were not sufficient to achieve adequate confounding adjustment, even with the use of automated data-adaptive PS method. Clinical information from the registry was needed to produce estimates similar to what was expected based on results from RCTs. At the same time, automated data-adaptive PS models performed as well as or better than the investigator-specified models across different databases and adjustment methods used.

It is worth noting that without detailed specification of variables by the researcher, automated data-adaptive PS was able to identify and include potential confounders in the model for the analysis for both registry-only and claims plus registry databases. This finding is consistent with prior studies of hdPS using claims databases^9,23–27^ and underscores the usefulness of the approach, especially in the absence of detailed pre-specification of confounders due to factors such as the need to conduct a large number of comparisons (e.g. monitoring and signal detection^28^), or when no previous assessment of potential confounders is available. At the same time, automated data-adaptive approach was not able to overcome the lack of information on patients’ high surgical risk status which was the most important confounding by indication in the current study.

While the registry is designed to collect clinical information for comparative effectiveness studies of the two procedures, adjustment using only registry information was not able to achieve estimates that adequately approximated those expected, and variables from claims data were needed to achieve maximum confounding adjustment. This supports the idea that claims databases potentially provide important information on confounders not captured in clinical or device registries. This is likely true for a few reasons. First, registries for specific diseases or devices tend to have sparse information on comorbidities that are not directly related but may affect receipt of treatment and outcomes (e.g., COPD or cancer for cardiovascular registries), whereas claims data capture all clinical encounters of patients regardless of the targeted diseases. Second, claims data can provide proxies of non-clinical health determinants, including use of services and therefore patients’ tendency to seek health care, their socioeconomic status, or frailty^29^. Automated data-adaptive approach may be especially useful in this context, as it can identify these proxies without investigators’ pre-specification. Other methods for identifying these health determinants from claims data using data mining techniques may further improve confounding control in these comparative studies.

Limitations of our study should be noted. First, to assess the performance of different adjustment approaches, we used the reported relative risk estimates from randomized clinical trials as a benchmark. However, results from randomized studies may differ from comparative effectiveness estimates obtained using routine care data due to differences in patient populations and possible effect measure modification. Second, the precision of our estimates was poor, due to the limited number of patients and events in the linked cohort, and we were thus unable to conduct meaningful statistical tests to compare the estimates. Third, the example study we used is for a particular comparative analysis and may have limited generalizability. While we expect combinations of claims database and registries with common characteristics to show similar tendencies in terms of the usefulness of agsPS for other comparative effectiveness research questions, additional studies with such combinations are needed to confirm our findings.

In summary, in a cardiovascular CER study with a high likelihood of confounding by indication, an automated data-adaptive approach for PS modeling performed well, with similar results seen for individual claims, registry, and claims plus registry databases. Although the c-statistics of propensity score models were similar across registry-based models with and without additional claims data components, neither was sufficient to adequately control confounding by indication by itself; both claims and registry variables were needed for better control. As expected, among the different data dimensions in the registry and claims data, the most important to control for confoudning by indication was high surgical risk staus data in the registry. Despite its potential to control for unmeasured confounding, the automated data adaptive approach itself was not sufficient to control strong confounding by indication when the information is lacking in the data source.

## Data Availability

The data used for the study are available to researchers through data use agreement with CMS as well as the Society for Vascular Surgery's Vascular Registry (SVS-VR). The corresponding author should be contacted for access to computing code.

